# Racial and Ethnic Disparities in Clinical Outcomes among Patients with Takotsubo Syndrome; A Nation-Wide Analysis

**DOI:** 10.1101/2024.02.18.24302960

**Authors:** Jamal Christopher Perry, Oluwasegun Matthew Akinti, Chukwuka Eneh, Henry Osarumme Aiwuyo, Charles Poluyi, Ukenenye Emmanuel, Esther Doudu, Henry Alberto Becerra, Mustafa Bilal Ozbay, Kibwey Roderick Peterkin, Rosy Thachil, Abdullah Khan

## Abstract

**Background:** Takotsubo syndrome (TTS), a stress-induced transient left ventricular dysfunction, remains poorly understood, with an estimated incidence of 1-2% among acute coronary syndrome cases. This study investigates racial and ethnic disparities in hospital outcomes and clinical characteristics of TTS.

**Methods:** We conducted a retrospective cohort study using the National Inpatient Sample data from 2016 to 2020, identifying TTS cases through validated ICD-10 codes. Statistical analysis was performed using Stata 18, with logistic regression models adjusting for confounders to identify disparities in outcomes.

**Results:** The study included 32,785 TTS hospitalizations; the majority were White (80.5%), followed by Black (6.7%) and Hispanic (5.8%) patients. Minority groups, mainly Black and Hispanic patients, were younger (average age 63) and predominantly from lower-income brackets, while paradoxical Asians had the highest income bracket. Notable findings include Black patients showed the highest rate of stroke (4.8%, OR 2.1, p = 0.003). The rate of cardiogenic shock was highest among Asians (11%, OR 2, p < 0.001). Mortality rates were elevated in Black (2%, OR 1.5, p < 0.001) and Asian populations (1.8%, OR 1.97, p < 0.001).

**Conclusion:** Significant racial and ethnic disparities exist in TTS outcomes, with minority groups having more in-hospital outcomes. These findings highlight the urgent need for targeted interventions and further research to reduce healthcare inequities in TTS management.

## Introduction

Takotsubo syndrome or cardiomyopathy (TTS), first described in Japan in 1990, manifests as transient left ventricular dysfunction triggered by emotional or physical stress, resembling a “Takotsubo” octopus trap^1.^ This condition predominantly affects postmenopausal women and mimics heart attack symptoms, though it occurs without coronary artery obstruction^1,2^. The underlying mechanisms of TTS are not fully understood. However, proposed theories include stress-induced increases in catecholamines, microvascular dysfunction, inflammation, estrogen deficiency, coronary vessel spasms, and myocardial stunning without obstructive coronary artery disease ^3^. TTS can have a favorable prognosis but can be associated with fatal complications ^2^. The real incidence of TTS remains unclear. It makes up for 1 to 2% of patients suspected of having acute coronary syndrome^3^.

TTS cardiomyopathy demonstrates an evident correlation with physical stressors (36%) more than emotional stressors (27.7%) ^6^. While studies have elucidated various aspects of this syndrome, there remains a paucity of research investigating potential racial and ethnic disparities among affected patients. Understanding such disparities is essential for optimizing patient care and treatment strategies and addressing healthcare inequities.

Our study focused on exploring racial and ethnic disparities in-hospital outcomes for patients diagnosed with Takotsubo Cardiomyopathy (TTS). Through a systematic analysis that included patient demographics and hospitalization metrics such as length of stay and total charges, and clinical outcomes, we offer a comprehensive view of the disparities within the healthcare system. Based on documented racial differences in outcomes for other cardiovascular conditions, we hypothesize that minority groups will have worse in-hospital cardiovascular outcomes compared to White patients with TTS.

## Methods

Using the National Inpatient Sample (NIS) Database Registry, we analyzed data from recent years 2017 to 2020.

### Database

The National Inpatient Sample (NIS) is part of the Healthcare Cost and Utilization Project (HCUP) and is maintained by the Agency for Healthcare Research and Quality (AHRQ) ^4^. The NIS contains information on all inpatient stays (not individual patients) in 48 states plus the District of Columbia, representing approximately 98% of the United States population, excluding rehabilitation and long-term acute care hospitals ^4^. Unweighted, it contains data from more than 7 million hospital stays each year, and weighted, it estimates more than 35 million hospitalizations nationally ^4^ observations in the NIS contain a primary diagnosis, up to 39 secondary diagnoses, and up to 25 procedure codes, depending on the year ^4^. All discharge diagnoses were identified using the International Classification of Diseases, 10th edition (ICD-10) codes ^5^. The AHRQ made these data available to the principal author via the HCUP.

### Statistical Analysis

National estimates were obtained using the discharge-level weight variable (DISCWT) provided by the HCUP. Weighted data were used for all statistical analyses. Missing values were excluded from the analysis. Categorical variables were compared using the Chi-square test and are described using frequency with percentages. Continuous variables were compared using the student’s t-test and are reported as mean (±SD) if their distribution was normal or compared using the Mann–Whitney U test and reported as median (interquartile range [IQR]) if their distribution was skewed. As part of our hypothesis-driven approach, we formulated and tested a null hypothesis asserting that there are no differences in outcomes of Takotsubo cardiomyopathy among racial and ethnic groups. Multivariate linear and logistic regression was used to compare hospital outcomes between groups, adjusting for potential confounders, which included age, Charlson comorbidity index, hospital bed size, teaching status, and insurance. Statistical analysis was performed using Stata version 18 (Stata Corp, College State, Texas). A p-value **≤** 0.05 was considered to indicate statistical significance for all analyses. Variables used in the regression model building were either selected from the dataset as provided variables or abstracted with the ICD-10 codes.

### Inclusion criteria and study variables

The study cohort consisted of all hospitalizations among individuals categorized as either white, Hispanic, Black, Asian, Native American Indian, and other races. Patients who had missing race or reported other race/ethnicity were included as others. We extracted the following independent variables: sociodemographic characteristics (including sex, age, mean annual income in zip code, and insurance type), hospital characteristics, comorbidities, Charlson comorbidity index, and outcomes (Tables 1 and 2). We used ICD-10 codes to identify principal/secondary diagnoses. To investigate disparities, we studied baseline characteristics and outcomes for TTS hospitalizations among all races (Supplemental Table S1).

**Table 1.**
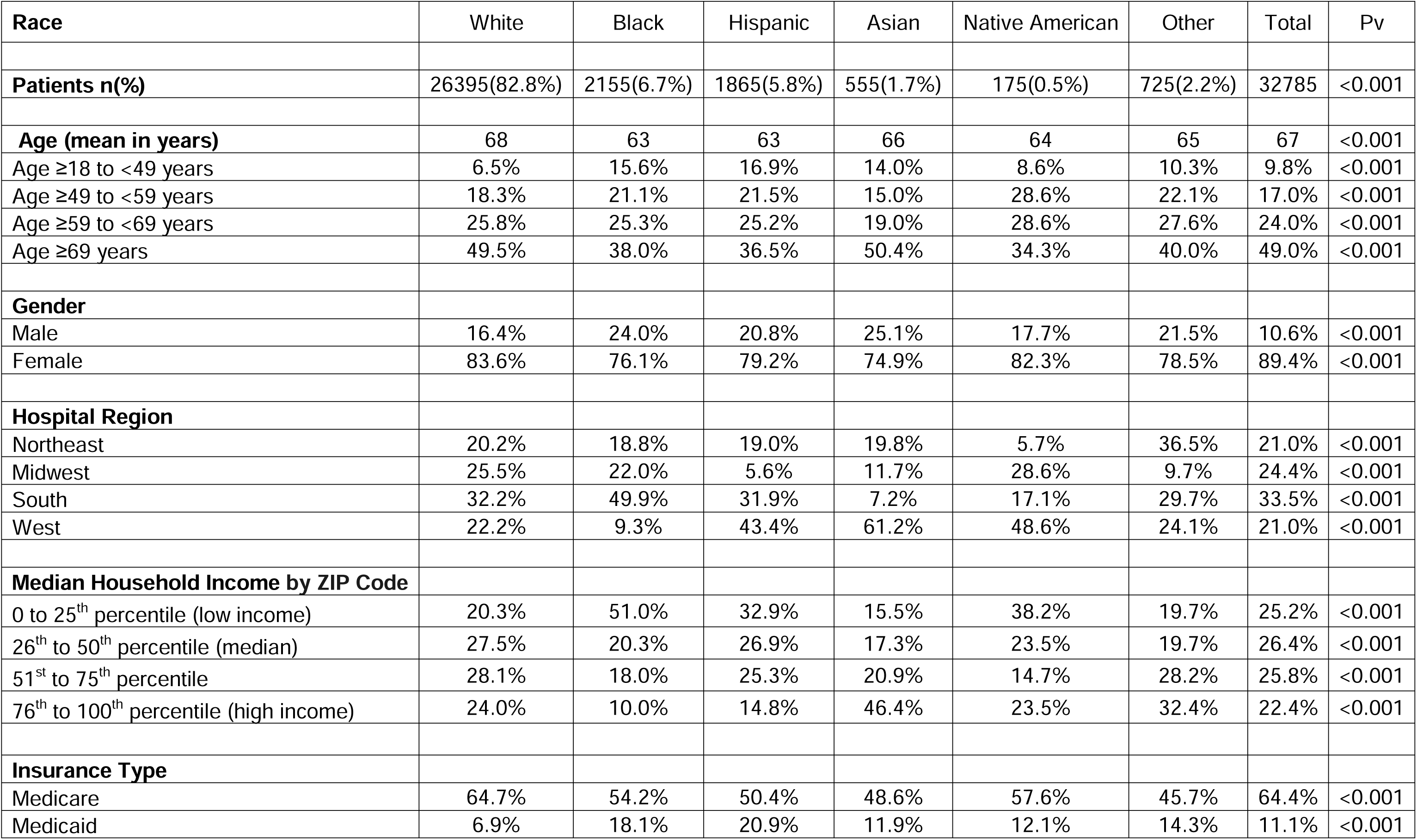
Patient Characteristics.

**Table 2.**
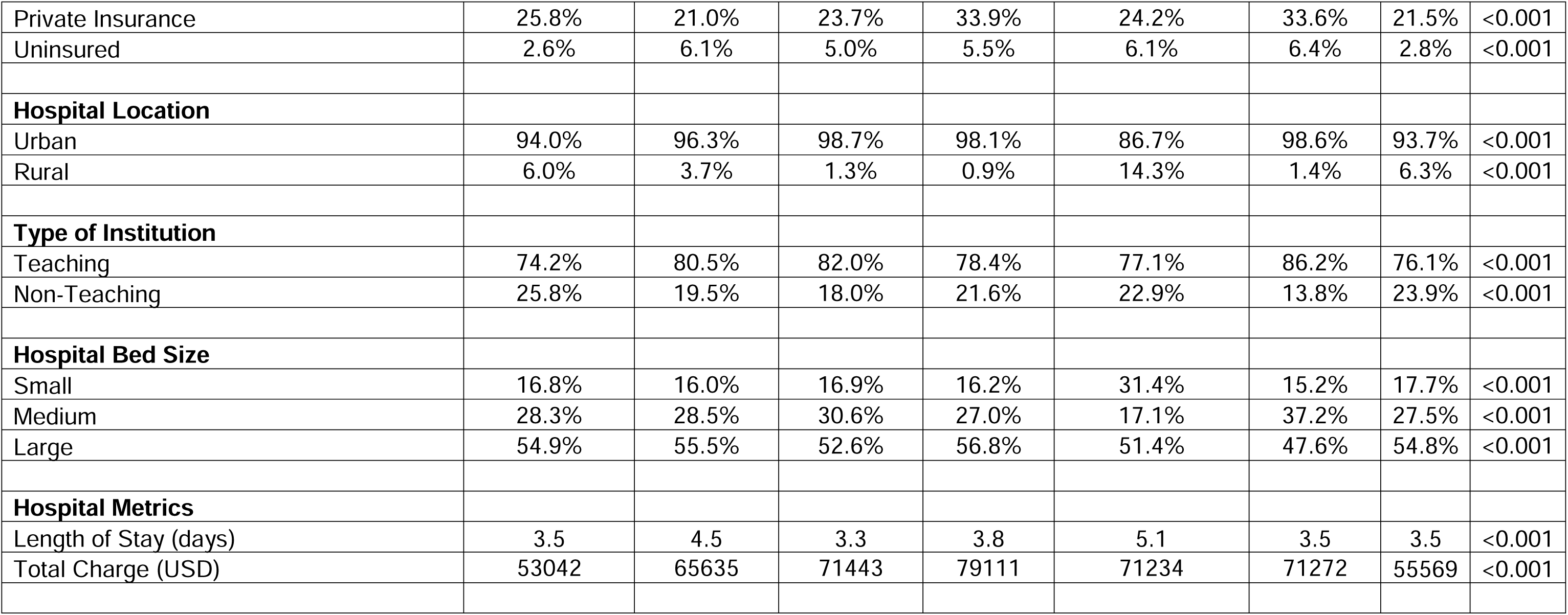

## Results

### Patient Demographics and Hospital Metrics

Among 32,785 hospitalizations with a principal diagnosis of TTS, demographic distributions were as follows: White patients comprised 26,395 (80.5%), Black patients 2,155 (6.7%), Hispanic patients 1,865 (5.8%), Native American Indians 175 (0.5%), Asian patients 555 (1.7%), and others 725 (2.2%). The average age was 67, with 89.4% of patients being female. However, among Black and Hispanic patients, the average age was 63. The primary hospital location was in the Southern region for 50% of the Black patient population, 32.15% of White patients, and 43% of Hispanic patients, compared to 48% of Native American Indians and 61.2% of Asian patient populations in the Midwest. Most patients were below the 50th percentile of income: 50% of Black patients, 38% of Native American Indians, and 32% of Hispanic patients were in the 0-25th percentile, compared to 52% of White patients and 66% of Asian patients, which were above the 50th percentile (Table 1 and Figure 1).

**Figure 1:**
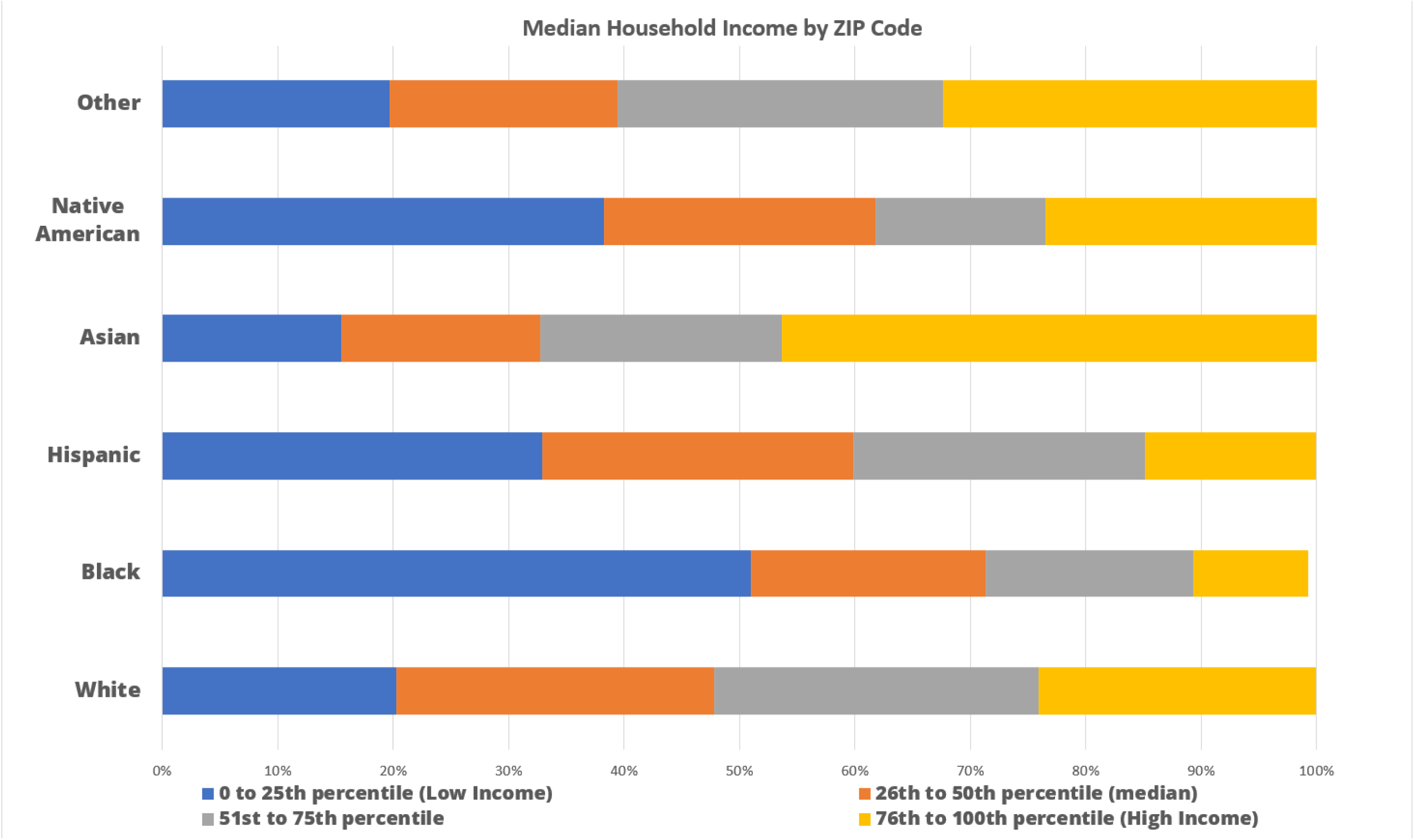
Median Household Income Distribution by Racial/Ethnic Group This figure illustrates the distribution of median household income across different racial/ethnic groups categorized by ZIP code income percentiles for the year 2020. The x-axis represents the percentage of households within each income percentile range, while the y-axis lists racial/ethnic groups. The colors represent different income percentile ranges: blue for the 0 to 25th percentile (Low Income), orange for the 26th to 50th percentile (Median Income), gray for the 51st to 75th percentile, and yellow for 76th to 100th percentile.

### Length of Stay (LOS) and Total Charges (TOTCHG)

The mean length of stay (LOS) among patients with TTS was 3.5 days. Black patients had a mean LOS of 4.5 days with a mean increase of 1.1 days (p-value <0.001, 95% CI 0.6 to 2.5), and Native American Indian patients had a mean LOS of 5.1 days with an increase of 1.75 days (p-value 0.045 95% CI 0.3-3.5), compared to LOS of 3.5 days for White patients with a mean decrease of 1.1 days (p-value 0.002, 95% CI −1.6 to −0.5). However, Asian patients had a mean LOS of 3.8 days with an increase of 0.4 days (p-value 0.17), and Hispanic patients had a mean LOS of 3.3 days (p-value 0.9), others with 3.5 LOS (p-value), which were not statistically significant. (Tables 1 and 3)

**Table 3.**

| Comorbidities | White | Black | Hispanic | Asian | Native American | Other |
| --- | --- | --- | --- | --- | --- | --- |
| Heart Failure | 34% | 36% | 29% | 36% | 37% | 32% |
| Mood Disorders | 2.4% | 4.9% | 2.7% | 1.8% | 5.7% | 4.1% |
| Smoker | 45% | 45% | 28% | 23% | 29% | 30% |
| Hyperlipidemia | 52.3% | 47.6% | 49.0% | 53.2% | 34.2% | 49.0% |
| Obesity | 11.8% | 14.1% | 14.8% | 5.9% | 15.7% | 9.6% |
| Hypertension | 38.3% | 40.8% | 43.7% | 38.7% | 31.4% | 37.9% |
| Diabetes Mellitus | 19.0% | 27.6% | 32.7% | 31.5% | 31.4% | 24.8% |
| Charleson Comorbidity Index: |  |  |  |  |  |  |
| 0 | 25.1% | 20.7% | 25.1% | 26.3% | 26.3% | 31.8% |
| 1 | 28.6% | 26.9% | 29.7% | 27.8% | 27.8% | 25.0% |
| 2 | 21.4% | 20.5% | 20.9% | 14.3% | 14.3% | 23.9% |
| ≥3 | 24.9% | 31.8% | 24.2% | 31.6% | 31.6% | 19.3% |
| Outcomes | White | Black | Hispanic | Asian | Native American | Other |
| Mortality | 1.6% | 2.0% | 0.5% | 1.8% | 2.9% | 4.1% |
| Pulmonary embolism | 1.4% | 2.5% | 1.7% | 0.6% | 1.0% | 3.7% |
| Stroke | 2.3% | 4.9% | 2.7% | 1.8% | 5.7% | 4.1% |
| Ventricular Arrhythmias | 4.6% | 5.8% | 4.3% | 2.7% | 8.5% | 7.6% |
| Cardiogenic shock | 6.3% | 8.3% | 8.8% | 11.9% | 8.6% | 7.7% |
| Cardiac arrest | 1.7% | 3.9% | 2.1% | 3.6% | 5.7% | 3.5% |
| Heart Failure with Preserved Ejection Fraction | 3.4% | 5.3% | 4.0% | 0.8% | 2.2% | 1.7% |

| Outcomes | White |  | Black |  | Hispanic |  | Asian |  | Native American |  | Other |  |
| --- | --- | --- | --- | --- | --- | --- | --- | --- | --- | --- | --- | --- |
|  | OR<sup>a</sup> (95% CI) | P<sub>v</sub> | OR (95% CI) | P<sub>v</sub> | OR<sup>a</sup> (95% CI) | P<sub>v</sub> | OR<sup>a</sup> (95% CI) | P<sub>v</sub> | OR<sup>a</sup> (95% CI) | P<sub>v</sub> | OR<sup>a</sup> (95% CI) | P<sub>v</sub> |
| **Pulmonary embolism** | 0.53(0.4-0.5) | <0.001 | 1.9(1.4-2.4) | <0.001 | 1.2 | 0.2 | 0.4 | 0.09 | 0.7 | 0.6 | 2.76(1.8-4.2) | <0.001 |
| **Mortality** | 0.67(0.6-0.8) | <0.001 | 1.5(1.2-1.7) | <0.001 | 1.4(1.1-1.6) | <0.001 | 1.97(1.5-2.5) | <0.001 | 1.2 | 0.4 | 1.8(1.4-2.3) | <0.001 |
| **Stroke** | 0.4(0.3-0.7) | 0.003 | 2.1(1.3-3.4) | 0.003 | 1.1 | 0.7 | 0.7 | 0.7 | 2.4 | 0.2 | 1.7 | 0.2 |
| **Ventricular Arrhythmias** | 0.8(0.7-0.9) | 0.04 | 1.2(1-1.3) | 0.046 | 0.9 | 0.7 | 1 | 0.8 | 1.2 | 0.4 | 1.3(1-1.7) | 0.04 |
| **Cardiogenic shock** | 0.7(0.6-0.8) | <0.001 | 1.35(1.2-1.5) | <0.001 | 1.46(1.2-1.7) | <0.001 | 2.(1.6-2.5) | <0.001 | 1.4 | 0.17 | 1.2 | 0.12 |
| **HFpEF** | 0.6(.41- 0.94) | 0.026 | 1.6(1.06 - 2.42) | 0.03 | 1.2(0.73 - 1.93) | 0.49 | 0.2(0.03 - 1.57) | 0.13 | 0.7(0.09 - 4.78) | 0.68 | 0.5(0.16 - 1.57) | 0.23 |
| **Cardiac arrest** | 0.43(0.2 - 0.7) | 0.002 | 2.3(1.3-3.9) | 0.002 | 1.2 | 0.56 | 2.1 | 0.7 | 3 | 0.2 | 2 | 0.18 |
| **Multivariate Linear Regression** |  |  |  |  |  |  |  |  |  |  |  |  |
| **Length of Stay** | -1.1(-1.6 to -0.5) | <0.001 | 1.1(0.48-1.6) | <0.001 | -0.03(-0.4-0.36) | 0.9 | 0.43(1.8-4.2) | 0.17 | 1.75(-0.0-3.5) | 0.045 | 0.4.(0.6-0.7) | 0.9 |
| **Total Charge** | -14442(-23931-4954) | <0.001 | 12019(1785-22252) | <0.02 | 15431(5497-25364) | 0.002 | 22858(3377-42339) | 0.02 | 15629(11045-42303) | 0.251 | 18950(1959-35939) | 0.029 |

HFpEF= Heart Failure with Preserved Ejection Fraction
OR =Odds Ratio
Pv= p-value

The mean total charge (TOTCHG) among patients with TTS was $55569. Black patients faced a mean TOTCHG of $65,635, with a mean increase of $12,019 (p = 0.02, 95% CI 1,785-22,252). Hispanic patients had a mean TOTCHG of $71,443, with a mean increase of $15,874 (p = 0.002, 95% CI 5,497-25,364). The highest mean TOTCHG was noted for Asian patients at $79,111, an increase of $23,542 (p < 0.02, 95% CI 3,377-42,339). Patients categorized as other races had a mean TOTCHG of $71,272, with an increase of $15,703 (p < 0.02, 95% CI 3,377-35,939). Conversely, Native American Indians and White patients had mean TOTCHGs of $71,234 and $53,042, respectively. Notably, White patients experienced a decrease of $14,442 from the mean (p < 0.001, 95% CI −23,931 to −4,954). (Tables 1 and 3)

### Heart Failure with preserved Ejection Fraction and Cardiogenic Shock

Heart Failure with preserved Ejection Fraction (HFpEF) rates were 3.4% among White patients (OR p-value 0.026 95% Cl 0.6 0.41-0.94) Native Americans was 2.2% (OR 0.7, p-value 0.68). Hispanic was 4% (OR 1.2, p-value 0.49), Asians 0.8% (0.2, p-value 0.13) compared to 5.3% among Black patients (OR 1.6, p-value 0.03, 95% CI 10.6 to 2.42).

Cardiogenic shock was reported in 8.25% of Black patients (OR 1.34, p-value <0.001, 95% CI 1.16 to 1.56), 8.8% of Hispanic patients (OR 1.4, p-value <0.001, 95% CI 1.2 to 1.7), and 11% of Asian patients (OR 2, p-value <0.001, 95% CI 1.5 to 2.5), compared with Native American Indian patients at 8.59% (p-value 0.172), other races at 7.6% (p-value 0.121), and White patients at 6.3% (OR 0.7, p-value <0.001, 95% CI 0.6 to 0.8).

### Pulmonary Embolism and Stroke

Stroke was noted in 4.8% among Black patients (OR 2.1, p-value 0.003, 95% CI 1.2 to 3.4) and 4.14% among other races (p-value <0.001), with Native American Indian patients at 5.7% (p-value 0.218), Asian patients at 1.8% (p-value 0.776), and Hispanic patients at 2.68% (p-value 0.720) compared to 2.39% among White patients (OR 0.4, p-value 0.003, 95% CI 0.3 to 0.7).

Pulmonary embolism was significant in 2.5% of Black patients (OR 1.8, p-value <0.001, 95% CI 1.4 to 2.4), other at 3.7% (OR 2.76, p-value <0.001, 95% CI 1.8-4.2), compared to 1.7% of Hispanic (p-value 0.227), 0.6% for Asian (p-value 0.094), 1% for Native American Indian (p-value 0.665), and 1.4% for White patients (OR 0.53, p-value <0.001, 95% CI 0.4-0.7).

### Mortality, Cardiac Arrest and Ventricular Arrhythmia

Inpatient mortality was 1.8% for Asian patients (OR 1.97, p-value <0.001, 95% CI 1.5 to 2.5), 0.5% for Hispanic patients (OR 1.38, p-value <0.001, 95% CI 1.17 to 1.63), 4.14% for other races (OR 1.8, p-value <0.001, 95% CI 1.6 to 2.1), and 2% for Black patients (OR 1.5, p-value <0.001, 95% CI 1.3 to 1.7) compared to 1.5% for White patients (OR 0.7, p-value <0.001, 95% CI 0.6 to 0.8) and Native American Indians (p-value 0.465).

Ventricular arrhythmia was only associated with the Black population at 5.8% (OR 1.2, p-value 0.041, 95% CI 1.1 to 1.3), while other races were not statistically significant.

Cardiac arrest was 3.9% in the Black population (OR 2.3, p-value 0.002, 95% CI 1.7 to 3.9), 2.1% for Hispanic (p-value 0.720), 3.6% for Asian (p-value 0.776), for Native American Indians (p-value 0.218), and for other (p-value 0.183) compared to 1.7% for White patients (OR 0.43, p-value 0.002, 95% CI 0.3 to 0.6). Outcome data for all races are presented in Tables 2 and 3.

## Discussion

We analyzed the national inpatient sample and noted potential disparities in patient characteristics and clinical outcomes among different racial and ethnic groups hospitalized with TTS.

We found that most TTS hospitalizations were among White patients, with Black, Hispanic, Native American Indians, and other minority groups making up a smaller portion of the total patient population. The average age was 67, and females accounted for 89% of the cases, aligning with existing research data ^6^. Our analysis also revealed that Black and Hispanic patients were, on average, younger at hospitalization (age 63), suggesting potential disparities in disease manifestation or healthcare access. Notably, 50% of the Black population with TTS fell within the lowest income quartile (0-25th percentile) (Figure 1). In our analysis, a notable distinction emerged within the Asian patient cohort affected by TTS, where 67.3% were identified in the higher income quartiles, particularly above the 51st percentile, indicating a majority within a high-income bracket.; the MASALA study explains that South Asian immigrants in the United States, have higher incomes largely due to selective immigration policies that favored professionals and those in demanded occupations, leading to a higher socioeconomic status. Despite higher socioeconomic status, they face an elevated risk of cardiovascular disease due to genetic predispositions, including higher levels of lipoprotein(a), which are associated with an increased risk of heart disease ^7^.

There were significant disparities in the length of hospital stays and hospital charges. Black and Native American patients experienced longer hospitalizations, a 2–3-day increase compared to the average. Asian patients encountered the highest total charges, while both Blacks and Hispanics incurred higher costs than their White counterparts (Figure 2).

**Figure 2:**
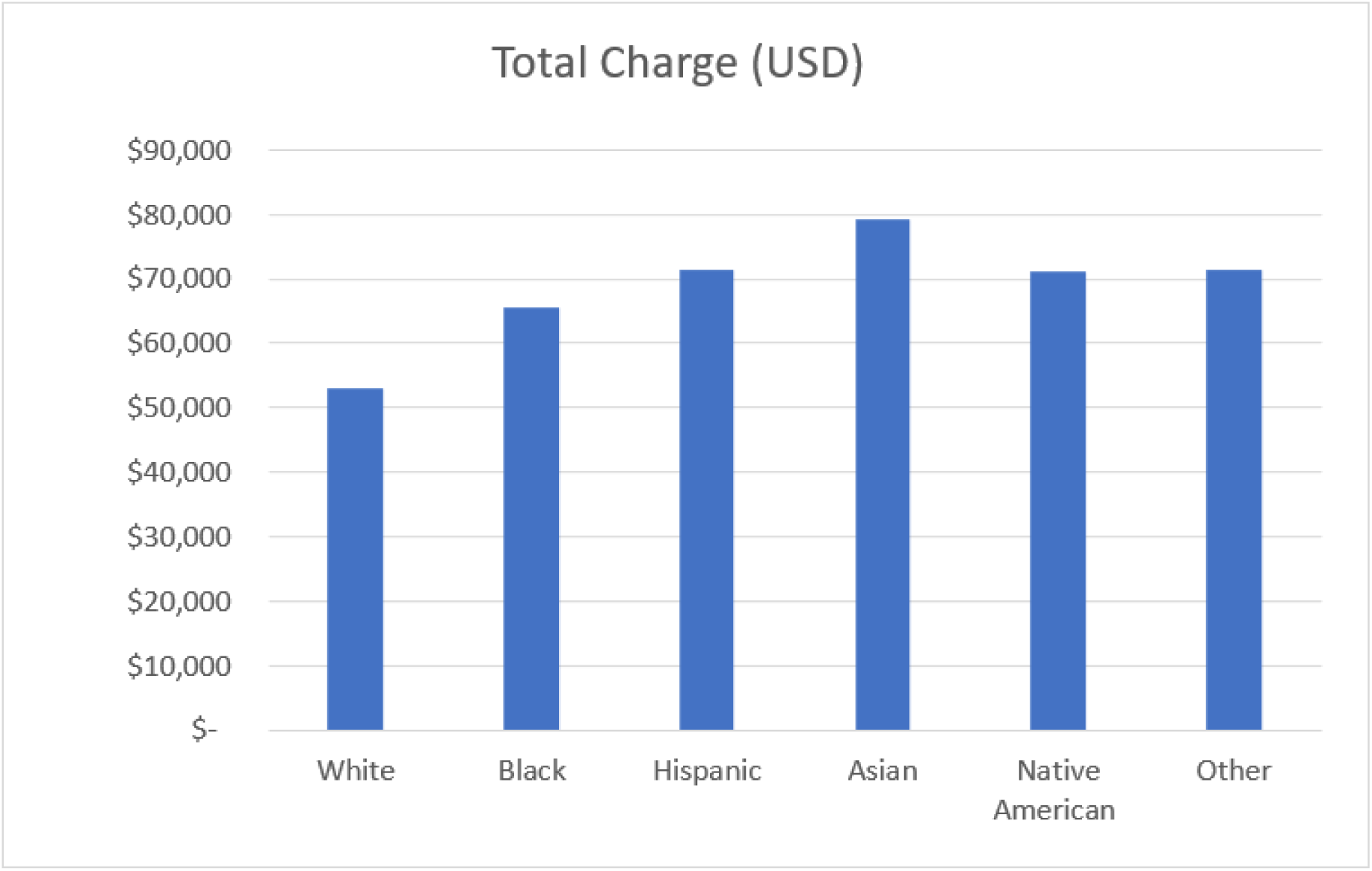
Total Hospital Charges by Racial and Ethnic Groups. This bar chart illustrates the total hospital charges incurred by patients, detailed on the y-axis in USD. Each bar represents a different racial or ethnic group. The groups are displayed on the x-axis and include White, Black, Hispanic, Asian, Native American, and Other.

These differences could reflect greater severity of disease presentation, delays in receiving care, or biases within healthcare facilities. The increased financial burden on Black and Hispanic patients highlights the urgent need for policy changes to ensure equitable healthcare costs.

Our findings indicated that Black, Asian, Hispanics, and Native American patients were associated with a higher in-hospital prevalence of comorbidities. Asian had the highest rate of cardiogenic shock followed by Black patients, who also experienced highest risk of HFpEF and ventricular arrhythmia.

This phenomenon may be attributed to significant disparities in cardiac care for Black individuals, encompassing prevention, invasive treatments, and outcomes ^8,9^. Notably, Black patients often face longer delays in receiving medications, undergoing revascularization, and obtaining surgical treatments for acute coronary syndrome, including extended door-to-balloon times ^9^. Genetic factors, such as the presence of the APOL1 and ADRB2 16R alleles in Black populations, might also play a crucial role in prompting more major cardiovascular outcomes ^10,11^. The etiology of cardiogenic shock appears to be multifactorial, suggesting a potential link between cardiogenic shock and other major cardiovascular complications prevalent among Black and Asian patients ^7,12, 13, 14^.

Pulmonary embolism was notably higher in Black patients (OR 1.8, p-value <0.001) and patients of other races (OR 2.3, p-value <0.001) compared to other populations. Stroke was the highest at 4.8% among Black hospitalized patients (OR 2.1, p-value 0.003) and 4.14% among other races (OR 1.7 p-value <0.001), significantly higher than in other races. In contrast, White patients showed no significant association (OR 0.4, p-value 0.003) with stroke. These outcomes suggest disparities may stem from underlying health conditions, differential access to preventative care, or variability in care standards.

Disparities in mortality rates were evident, with the highest inpatient risk of mortality observed in Asian (OR 1.97, p <0.001) and other race populations (OR 1.8, p <0.001), followed by 2% of Black patients (OR 1.5, p <0.001). Black patients also demonstrated a notable association with cardiac arrests (OR 2.3, p-value 0.002) compared to White patients, who showed no significant mortality association (OR 0.6, p <0.001). It was noted that the Black population had more major cardiovascular outcomes collectively compared to other races, which is consistent with literature ^15^.

Additionally, there is a hypothesis that chronic medical and socioeconomic stressors, which disproportionately affect specific minority racial groups such as Black communities, may influence the severity of Takotsubo syndrome outcomes ^16^. It is proposed that the more adverse outcomes observed in these populations could be partially explained by elevated levels of chronic stress, potentially leading to increased catecholamine production ^1^. This hypothesis is supported by existing research indicating that the convergence of medical and socioeconomic challenges is more prevalent in certain minority groups and may significantly contribute to the pathophysiology of Takotsubo cardiomyopathy ^16,1^.

In summary, in our analysis, we observed that Asian patients with higher socioeconomic status experienced a more significant association with cardiogenic shock and mortality, suggesting that genetic factors may play a more important role in cardiovascular outcomes in this population. Conversely, for Black and Hispanic patients, socioeconomic status cannot be ignored as a crucial determinant of health outcomes, consistent with existing literature^1,16,17^. These observations underscore the complexity highlighted by recent cardiovascular research, which argues that both genetic factors and social determinants of health are integral to understanding disease outcomes^18^. This aligns with the emerging consensus that a multifaceted approach is essential for managing TTS, where neither genetic nor socioeconomic influences can be considered in isolation.

### Limitations

The study faced several limitations. The reliance on an administrative database dependent on ICD10-CM codes, where incorrect coding could influence findings, was a primary concern. The lack of randomization was addressed using univariate and multivariate analysis. The strength of our study lies in the power of the sample size; we utilized a large population database, thus improving the validity of our findings. However, large databases can produce statistically significant but not clinically meaningful results, potentially overestimating minor differences. Our prevalence calculations used the total number of in-hospital TTS diagnoses from the NIS. We recognize limitations such as potential coding inaccuracies, missing data, the inclusion of selected hospitals, lack of longitudinal follow-up, and exclusion of outpatient data.

## Conclusion

This study sheds light on significant racial and ethnic disparities in Takotsubo Syndrome outcomes. Despite TTS’s higher in-hospital prevalence among White patients, patients from Black, Asian, and other racial and ethnic backgrounds face a greater risk of severe conditions, such as cardiogenic shock, and other major cardiovascular outcomes, especially among Black patients. These findings underscore the critical impact of socioeconomic factors, genetics, and healthcare access on patient outcomes, suggesting that disparities in these areas may contribute to delayed diagnosis, treatment, and increased complications. Isolating the effects of genetics from socioeconomic influences remains challenging, particularly as higher income in Asian populations did not correlate with better outcomes. Future studies incorporating genetic analyses may provide deeper insights into these complex interactions. Further research and interventions are needed to address these disparities, focusing on understanding and overcoming the socioeconomic and healthcare barriers that minority groups face.

## Data Availability

All data produced in the present work are contained in the manuscript and was sourced from NIS Database Documentation. Agency for Healthcare Research and Quality. https://www.hcup-us.ahrq.gov/db/nation/nis/nisdbdocumentation.jsp

## Authors and Contributions

- **Dr. Jamal Christopher Perry, MD** (Corresponding Author, First Author)

○ **Email**:
○ **Contributions**: Data Analysis, Data Collection, Software, Methodology, Writing - Original Draft, Review & Editing
- **Oluwasegun Matthew Akinti, MD**

○ **Contributions**: Writing - Original Draft, Review & Editing
- **Chukwuka Eneh, MD**

○ **Email**:
○ **Contributions**: Data Analysis, Software, Methodology, Writing - Original Draft, Review & Editing
- **Henry Osarumme Aiwuyo, MD**

○ **Email**:
○ **Contributions**: Writing - Original Draft, Review & Editing
- **Charles Poluyi, MD**

○ **Contributions**: Review & Editing
- **Ukenenye Emmanuel**

○ **Contributions**: Review & Editing
- **Esther Doudu**

○ **Contributions**: Review & Editing
- **Henry Alberto Becerra, MD**

○ **Contributions**: Review & Editing
- **Kibwey Roderick Peterkin, MD**

○ **Contributions**: Review & Editing
- **Mustafa Bilal Ozbay, MD**

○ **Contributions**: Review & Editing
- **Rosy Thachil, MD**

○ **Contributions**: Review & Editing
- **Abdullah Khan, MD**

○ **Contributions**: Supervision, Review & Editing

## Declaration of Interests

The authors declare that they have no known competing financial interests or personal relationships that could have appeared to influence the work reported in this paper.

## Disclosure Statement

### Disclosure

The authors declare that they have no competing financial interests or personal relationships that could have appeared to influence the work reported in this manuscript.

## Funding Sources

### Funding

This research did not receive any specific grant from funding agencies in the public, commercial, or not-for-profit sectors.

## Patient Consent

**The authors confirm that patient consent is not applicable to this article.** Research utilizing the National Inpatient Sample (NIS) data generally does not require Institutional Review Board (IRB) approval. This is due to the data being de-identified and made publicly available, which poses minimal risk to subjects. Furthermore, adherence to HIPAA regulations ensures the protection of patient privacy, thereby diminishing the necessity for IRB oversight in this context.

## Notes

### Competing Interest Statement

The authors have declared no competing interest.

### Funding Statement

No Fundings of any sort

### Author Declarations

NIS Database Documentation. Agency for Healthcare Research and Quality. https://www.hcup-us.ahrq.gov/db/nation/nis/nisdbdocumentation.jsp

### Summary of Updates

Figures 2 and 4 were removed Discussion was expanded References were added

