## Supplementary Table 1 for "Racial and Ethnic Disparities in Clinical Outcomes among Patients with Takotsubo Syndrome; A Nation-Wide Analysis"

**Supplemental Table S1**

| **Variables** | **ICD 10 Codes** |
| --- | --- |
| **Takotsubo Syndrome or Cardiomyopathy** | I5181 |
| **z** |  |
| **Hyperlipidemia** | E782, E784 |
| **Tobacco Use** | Z87891, F17x |
| **Mood Disorders include:**  1. Manic episode  2.bipolar disorder  3. Depressive episode  4. Major depressive disorder  5. Persistent mood disorders  6. Unspecified mood disorder | [F30](https://www.icd10data.com/ICD10CM/Codes/F01-F99/F30-F39/F30-)  [F31](https://www.icd10data.com/ICD10CM/Codes/F01-F99/F30-F39/F31-)  [F32](https://www.icd10data.com/ICD10CM/Codes/F01-F99/F30-F39/F32-)  [F33](https://www.icd10data.com/ICD10CM/Codes/F01-F99/F30-F39/F33-)  [F34](https://www.icd10data.com/ICD10CM/Codes/F01-F99/F30-F39/F34-)  [F39](https://www.icd10data.com/ICD10CM/Codes/F01-F99/F30-F39/F39-) |
| **Diabetes Mellitus** | E11, E13 |
| **Diastolic Heart Failure** | 1503* |
| **Obesity** | E66 |
| **Congestive Heart Failure** | I50x |
| **Chronic Kidney Disease** | N18x |
| **Hypertension** | I10 |
| **Stroke** | I60x, I61x, I62x , I69x, I63x, G46x, G45x, I6781, I6782, I97810, I97811, I97820, I97821 |
| **Ventricular Arrhythmia** (tachycardia and fibrillation) | I4901, I4902 |
| **Cardiogenic Shock** | R570 |
| **Cardiac arrest** | 146 |
| **Pulmonary Embolism** | I26 |
